# The Unmet Needs of People with Parkinson’s Disease and How They Relate to Current Late-Stage Clinical Trials

**DOI:** 10.1101/2022.06.28.22276886

**Authors:** Jasmine Moon, Monica Javidnia

## Abstract

**Objective:** To identify the symptoms and unmet needs of people with Parkinson’s Disease and characterize Phase 2-3 and Phase 3 clinical trials.

**Methods:** We surveyed people with Parkinson’s Disease to determine their symptoms, pharmacological and non-pharmacological treatments, side effects, and insufficiently managed or unmanaged symptoms, and wishes for Parkinson’s Disease research. We identified clinical trials through clinicaltrials.gov.

**Results:** Twenty-eight people with Parkinson’s Disease completed the study (22 survey, six interview), and 38 clinical trials were characterized. The majority of participants’ desires for clinical development and outcomes of clinical trials characterized targeted motor symptoms.

**Conclusions:** Despite numerous therapeutics available, motor symptoms continue to be a top priority for people with Parkinson’s Disease and focus for drug development.

All relevant ethical guidelines have been followed, all necessary IRB and/or ethics committee approvals have been obtained, all necessary patient/participant consent has been obtained and the appropriate institutional forms archived.

## Introduction

Parkinson’s disease (PD) is the fastest growing neurodegenerative disorder,^1^ characterized by motor and non-motor symptoms, including, but not limited to, tremor, bradykinesia, rigidity, postural instability, cognitive deficits, constipation, sleep disturbances, genitourinary dysfunction, and mood disorders.^2^ There are numerous pharmacological treatment options to address PD symptoms.^3,4^ However, due to treatment complications (e.g., levodopa-induced dyskinesia), symptoms that are insufficiently managed by current pharmacotherapies (e.g., cognition), and the lack of a disease-modifying treatment, there are continued efforts in PD drug development toward addressing therapeutic shortcomings. A number of experimental interventions are being studied preclinically and in various phases of clinical development.^5^ We sought to survey people with PD to understand their symptoms, unmet needs, and what they would like drug developers to focus on. Additionally, we characterized the symptoms targeted by current Phase 2-3 and Phase 3 PD trials.

## Methods and Procedures

### Study participants

We recruited English-speaking adults with PD, globally. Participants could either respond to questions via interview (US only) or survey (open globally). The study was approved by the University of Rochester Research Subjects Review Board, study number STUDY00004151.

### Survey questions

Participants were presented with the following questions, with the option to skip any question they did not wish to answer. Demographics: year of birth, year of PD diagnosis, sex, country you currently live in. Age and time with PD were derived from birth year and diagnosis year, respectively. The survey questions also included:

1. Parkinson’s disease can be associated with many symptoms such as tremor, constipation, fatigue, problems with balance, memory, mood, sleep, and more. These symptoms can differ from person to person. Would you describe some of the most bothersome symptoms you experience?
2. What medications are you taking for your Parkinson’s disease symptoms? These can be for the motor (movement-related) or non-motor symptoms.
3. Do you experience any side effects from the medications you take? If so, please state the side effect(s) and medication(s) they are associated with.
4. Aside from medication, what do you do to alleviate your symptoms? Examples include yoga, massage, support groups, etc.
5. Which one of your symptoms do you feel are insufficiently managed, meaning either you’re not getting treated for that symptom, or you are getting treatment, but it’s not really working that well for you?
6. What are some symptoms you wish researchers developing treatments for Parkinson’s disease would focus on?

Lastly, participants had an opportunity to provide any additional comments. Those who participated via interview were asked similarly-phrased questions.

### Review of clinical development for Parkinson’s disease

Clinical trials for PD therapeutics were identified through a review of interventional, Phase 2-3 and Phase 3 clinical trials for PD posted to clinicaltrials.gov (downloaded June 9, 2020). Data selected were from trials with the following recruitment statuses: not yet recruiting, recruiting, enrolling by invitation, active not recruiting (42 total studies). Four trials (NCT04287543, NCT04265209, NCT04193527, NCT03042416) were excluded as their trial aims and outcomes were outside of the scope of this manuscript.

## Results

### Participant Characteristics

Participant demographics, question response rates, and number of words and characters typed in survey responses are detailed in Table 1. Twenty-eight study participants were recruited, six via interview (21.4%) and 22 via survey (78.6%). Three questions (symptoms, medications, and wishes for PD research) had 100% response rates.

**Table 1.** Study participant demographics and question completion rate.

| **Participant demographics, n=28** |  |
| --- | --- |
| Sex, n (%) |  |
| Female | 15 (51.7%) |
| Male | 13 (44.8%) |
| Age, average ± standard deviation (range) | 60.3 ± 11.8 (40-77) |
| Years since diagnosis, average ± standard deviation (range) | 5.6 ± 6.9 (0-33) |
| Location, n (%) |  |
| United States | 20 (71.4%) |
| Non-United States | 8 (28.6%) |
| Overall response rate, n (%) |  |
| Symptoms | 28 (100%) |
| Medications | 28 (100%) |
| Medication side effects* | 25 (89.3%) |
| Symptoms unmanaged or insufficiently-managed | 26 (92.9%) |
| Non-pharmacological interventions* | 27 (98.4%) |
| Wishes for Parkinson's disease research | 28 (100%) |
| Survey response rate, n (%); word and character average (range) |  |
| Symptoms | 22, (100%) |
| Words, Characters | 10.1 (1-61), 56.4 (6-276) |
| Medications | 22, (100%) |
| Words, Characters | 5.0 (1-15), 33.5 (7-93) |
| Medication side effects | 20, (90.9%) |
| Words, Characters | 8.1 (1-41), 37.0 (2-163) |
| Symptoms unmanaged or insufficiently-managed | 20 (90.9%) |
| Words, Characters | 5.3 (1-34), 27.9 (3-126) |
| Non-pharmacological interventions | 22, (100%) |
| Words, Characters | 8.2 (1-34), 44.3 (4-185) |
| Wishes for Parkinson's disease research | 22, (100%) |
| Words, Characters | 7.8 (1-39), 38.4 (6-167) |

\*Question was not asked of one interview participant

### Pharmacological and non-pharmacological treatments

Participants reported an average of 2.6 drugs or supplements (range 0-7, 74 total), with 23 (82.1%) participants reporting use of levodopa and 55 of the 74 (72.6%) drugs/supplements reported targeting motor symptoms (one off-label for tremor). Additional reported substances addressed the non-motor PD symptoms pain/dystonia (n=4, 14.3%), sleep (n=3, 10.7%), anxiety (n=2, 7.1%), depression (n=2, 7.1%), constipation (n=1, 3.6%), fatigue (n=1, 3.6%), hypotension (n=1, 3.6%), and urinary dysfunction (n=1, 3.6%). Two participants reported participation in clinical trials and the use of investigational compounds, and one reported vitamin supplementation. Only one drug was not associated with PD symptoms.

Twenty-five total side effects in 14 categories were reported, ranging 0-3 per participant. Nine participants (36%) reported that they did not experience any side effects associated with their medications. The three most common were upper gastrointestinal side effects (n=6, 24.0%), fatigue/tiredness (n=5, 20.0%) and dyskinesia (n=4, 16.0%). No other side effect or complication was reported by more than one participant.

Among non-pharmacological approaches to addressing PD symptoms (average 2-9, range 0-10), exercise was the most popular of the 76 entries, with 21 of 26 respondents (80.8%) reporting some form of physical activity. Of those who reported a specific type of exercise, seven (26.9%) reported yoga/meditation/stretching, five (19.2%) reported boxing, four (15.4%) reported biking, and four (15.4%) reported walking/jogging/running. Of the top non-exercise approaches, five reported a support group or engaging the PD community (19.2%), four reported receiving massages (15.4%), and three received physical therapy (11.5%). Only one participant (3.8%) reported the use of vitamins, though this may have been due to the phrasing of the study question and examples provided.

### Symptoms

Participants reported an average of 3.4 symptoms (range 1-8, 96 total), with a fairly even balance overall between motor and nonmotor symptoms. We curated the entries into 28 symptoms or symptom categories (Figure 1A): tremor (n=15, 53.6\%), balance (n=11, 39.3\%), sleep (n=10, 35.7%), rigidity (n=9, 32.1%), fatigue (n=7, 25.0%), cognition (n=6, 21.4%), constipation (n=5, 17.9%), pain (n=4, 14.3%), upper gastrointestinal - nausea, appetite changes, (n=4, 14.3%), anxiety (n=3, 10.7%), dystonia (n=3, 10.7%), bradykinesia (n=2, 7.1%), dexterity (n=2, 7.1%), and hallucinations (n=2, 7.1%). Other symptoms were each reported once.

**Figure 1.**
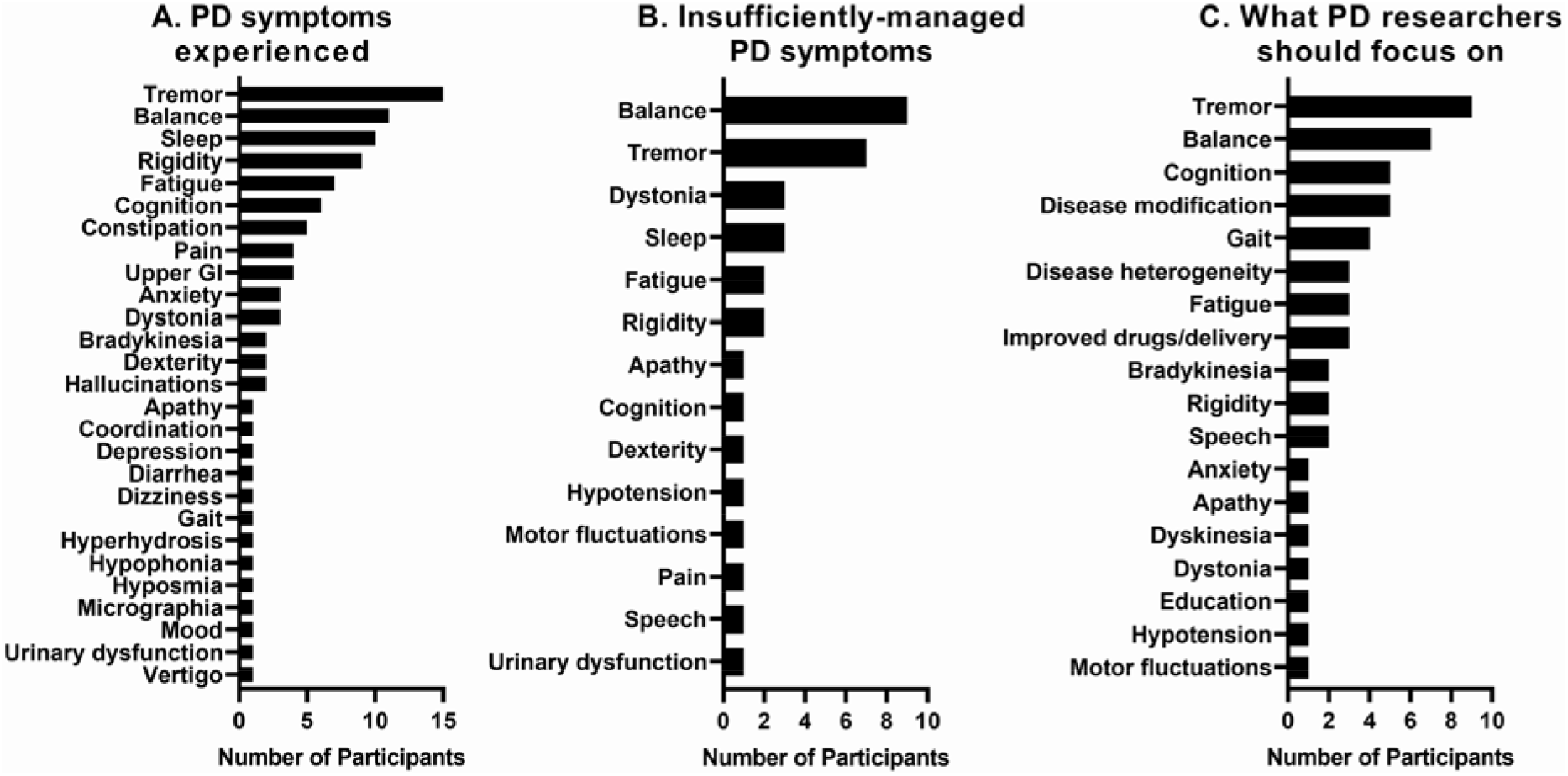
Distribution of participant responses. A. PD symptoms experienced, n=28 respondents. B. Unmanaged or insufficiently-managed PD symptoms, n=26 respondents. C. Wishes for PD research, n=28 respondents.

Thirty-four total unmanaged symptoms were provided (average 1.3, range 0-4), displayed in Figure 1B. The top two symptoms participants felt were unmanaged were motor-related: balance (n=9, 34.6%) and tremor (n=7, 26.9%). Three participants reported dystonia (11.5%), three reported sleep (11.5%), two reported fatigue (7.7%), and two reported rigidity (7.7%). Thought the predominance of motor symptoms was surprising, we did expect balance to be the most prevalent unmanaged symptom, as it is largely unresponsive to current PD pharmacotherapies. Only two participants (7.7%) reported they did not have PD symptoms that they feel are insufficiently managed, and other unmanaged symptoms were reported by one participant each (3.8%).

### Wishes for Researchers to Focus on

Among the 52 ‘wishes’ (average 1.9, range 1-4), treatments addressing tremor were the top wish, reported by nine participants (32.1\%), followed by balance (n=7, 25.0%), cognition (n=5, 17.9%), disease modifiers (n=5, 17.9%), gait (n=4, 14.3%), disease heterogeneity (including sex differences) (n=3, 10.7%), fatigue (n=3, 10.7%), improved drugs (rather than additional drugs) or drug delivery (n=3, 10.7%), bradykinesia (n=2, 7.1%), rigidity (n=2, 7.1\%), and speech (n=2, 7.1%), with several additional items reported by one participant each (Figure 1C).

### Overlap in Reporting

We quantified items reported in the symptoms, unmanaged symptoms, side effects/complications, and ‘wishes’ questions to determine whether there was an overlap in items reported by individual participants. No items spanned all four questions; however, there 29 instances when items were reported in two questions (1.0 average, 0-4), and 17 across three questions (0.6 average, 0-2). For symptoms that appeared across two questions (Supplemental Figure 1A), the top symptoms were fatigue (n=5), tremor (n=5), balance (n=3), sleep (n=3), dystonia (n=2), and rigidity (n=2). Nine additional symptoms overlapped once. For symptoms that appeared across three questions (Supplemental Figure 1B), the top overlapping symptoms were balance (n=7) and tremor (n=5), with five additional symptoms that overlapped three questions once each.

### Phase 2-3 and 3 trials

Of the 38 studies identified, two were testing aerobic exercise as an intervention. Sixteen trials (42.1%) targeted motor symptoms, fluctuations, or levodopa-induced dyskinesia, six trials (15.8%) tested potential disease-modifying treatments, three trials (7.9%) were testing ampreloxetine for symptomatic neurogenic orthostatic hypotension, two trials (5.3%) focused on pain, two trials (5.3%) were for falls or postural instability, and another two (5.3%) were for non-motor symptoms broadly. The five remaining trials were testing interventions for anxiety, depression, impulse-control disorders, quality of life, and urinary symptoms (one trial, each).

## Discussion

Since the introduction of oral levodopa in 1967 for PD6, numerous motor therapeutics have been introduced, including PD anticholinergics, dopamine receptor agonists, monoamine oxidase-B inhibitors, and others. Our findings suggest despite the numerous treatments available for motor symptoms, improved drugs that better address symptoms and complications are still greatly needed. Another top concern across PD symptoms reported, PD symptoms insufficiently managed, and what participants felt PD resources should focus on was balance, a symptom for which pharmacotherapies do not currently exist. Balance and tremor are also, repeatedly, the top symptoms reported by patients, further emphasizing the need to develop drugs that address such symptoms.

There were some shortcomings to this study. The survey was launched in the early stages of the COVID-19 pandemic, and, expectedly so, the response rate was not as anticipated. As such, we were unable to further explore potential sex differences in responses or factors such as age or geography. Additionally, the number of non-motor symptoms reported was lower than expected.

## Conclusions

This is the first study to survey people with PD in order to look at three different factors: patients’ symptoms, unmet needs, and what patients would like drug developers to focus on. Our main finding that most of the PD symptoms that are insufficiently addressed for people with Parkinson’s Disease were regarding motor symptoms, specifically balance. This finding is significant as it highlights the need for the further development of drugs that address such movement symptoms. In future studies, we will include a checklist option of all known PD symptoms as well as space for participants to include symptoms not listed.

## Data Availability

All data produced in the present work are contained in the manuscript.

