## Supplemental Figure 1 for "The Unmet Needs of People with Parkinson’s Disease and How They Relate to Current Late-Stage Clinical Trials"

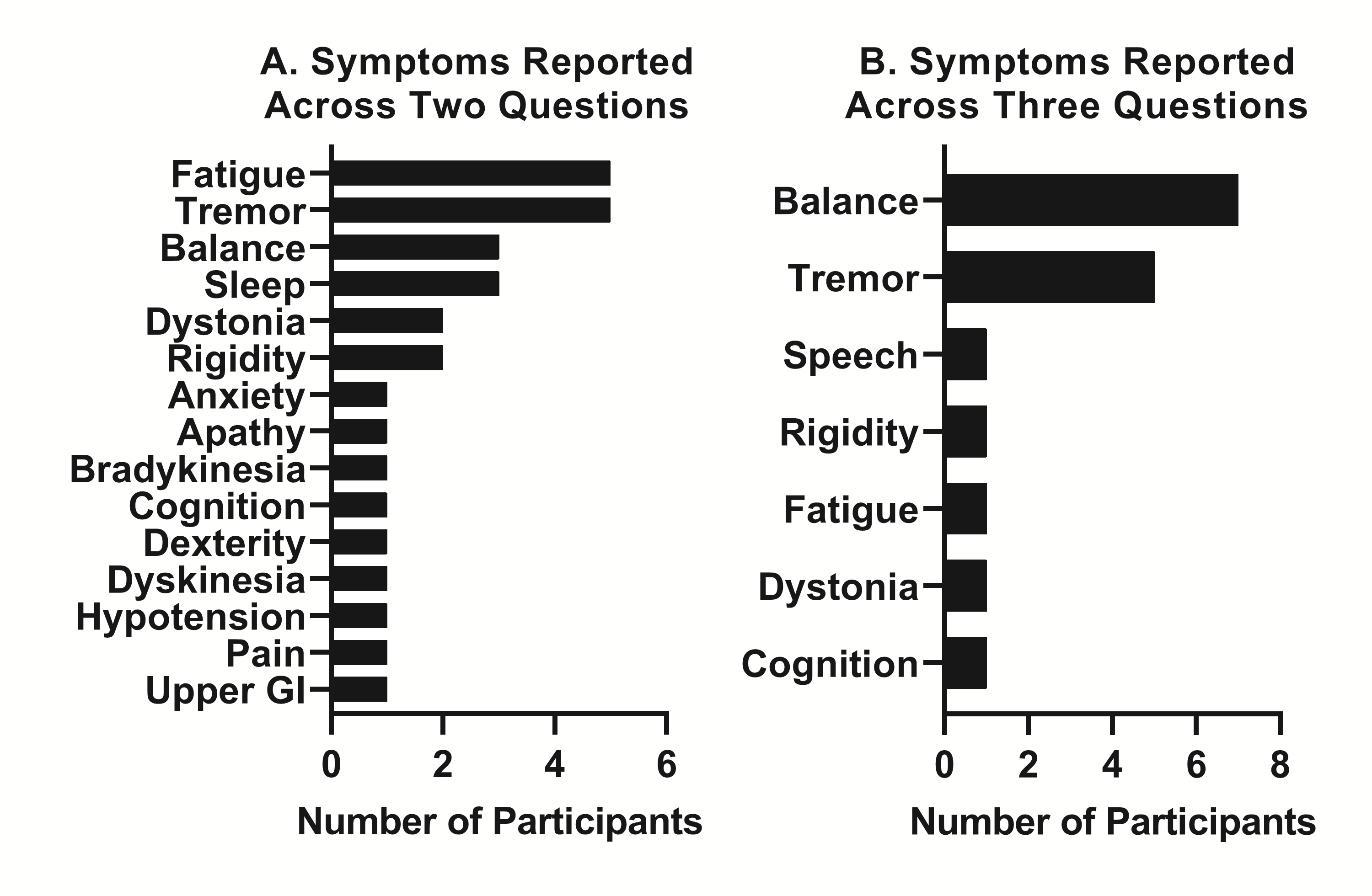

Supplemental Figure 1. Symptom and side effect reporting across multiple questions.

A. Symptoms and side effects that were reported across two questions. B. Symptoms and side effects reported across three questions.
